# Exploring the association between cannabis and opioid use among adults in Washington State: Estimates from a Cross-Sectional Survey

**DOI:** 10.1101/2023.11.27.23299105

**Authors:** Eslam Abousamra, Gabriel Andres, Alyson J Litman

## Abstract

**Introduction:** Cannabis and opioids are commonly used for pain management. However, studies examining the association between use of both substances conflict. Furthermore, disruptions to healthcare services and access due to COVID-19 may have affected opioid use in favor of cannabis because of accessibility. Our objective was to examine associations between cannabis and opioid use in Washington State (WA), COVID-19’s impact on these associations, and the association between frequency of cannabis use and misuse of opioids.

**Methods:** We pooled cross-sectional data from 2019 and 2021 WA Behavior Risk Factors Surveillance Systems surveys to examine associations between cannabis use in the past 30 days and prescription or non-prescription opioid use and misuse during the past 12 months among adults 18 and older. Survey-weighted adjusted prevalence ratios were estimated using multivariable Poisson regression. A proportion test and adjusted trend test were used to examine trends between cannabis use frequency (no use, 1-5 days, 6-9 days, 10-29 days, and 30 days) and opioid misuse among those reported opioid use.

**Results:** Among 25,540 participants, the prevalence of opioid use was 19.2% among those who used cannabis and 13.8% among those who did not use cannabis (aPR=1.32; 95%CI: 1.20,1.46). COVID-19 did not modify the association between cannabis and opioid use. Among those who used opioids (n=2,168), daily cannabis use was associated with higher opioid misuse prevalence (aPR=2.64; 95% CI: 1.71,4.10), though less than daily use was not.

**Conclusion:** Cannabis and opioid use were positively associated. Our findings emphasize the need for cautious policy decisions on cannabis regulations. Promoting cannabis use may not correlate with a reduction in other substance misuse. Further research is warranted to explore patterns of cannabis and opioid misuse.

## INTRODUCTION

In the United States (US), deaths from opioids have quintupled since 1999. In just a one-year span (2019 to 2020), deaths from drug overdose increased by 30%, with nearly

69,000 people dying of an opioid-related overdose in 2020.^1^ Similar trends have been observed in Washington State (WA).^2^ Opioid misuse has created a financial burden for WA, with a combined cost of opioid use disorder (OUD) and fatal opioid overdose estimated at $23.6 billion in 2017.^3^ Some factors that have contributed to the opioid epidemic in WA have been well studied, and are currently monitored, such as the increase in the prescription of opioids by healthcare professionals.^2^ However, some factors and how they influence opioid use are not well understood. One of those factors is cannabis use.

Recent studies exploring the association between cannabis use on opioid misuse are conflicting.^4,5,6^ In three recent studies exploring the association between cannabis and opioid use, two observed a positive association and one observed an inverse association.^4,5,6^ The inverse association was observed in a clinical population in treatment for OUD in Ontario, Canada.^4^ This study found no association between cannabis use and opioid use. However, patients experiencing cannabis-related side effects had lower odds of opioid use compared to those with no cannabis-related side effects (OR = 0.66,95% CI 0.52, 0.84, p=0.001).^4^ Even so, these findings were limited to individuals with OUD and therefore were not generalizable to the general population.^4^ A positive association between cannabis use and non-medical opioid use was observed in a National Epidemiological Survey representing the non-institutionalized US adult population.^5^ In a recent cross-sectional study of adults in the United States with chronic pain, higher levels of anxiety and depression were associated with concurrent use of opioids and cannabis.^7^ However, neither study examined whether opioid use varies depending on the frequency of cannabis use, limiting inferences regarding causality and dose-response relationship between cannabis and opioid use.^4,5^

The opioid epidemic in WA is one of the state’s forefront health issues and identifying solutions will be difficult without a complete understanding of the factors influencing opioid-related outcomes. Cannabis use has been implicated as both a positive and negative influence on opioid use in prior literature, however, no studies have investigated this association in the aftermath of COVID-19.^4,5,6^ The additional stressors of the COVID-19 pandemic, such as increased stress and anxiety, limited access to pain-management and/or mental health providers, and disruptions in healthcare services, may have led individuals to use cannabis as a means of self-medication for various physical and emotional distress, potentially impacting the use of opioids.^8^

Furthermore, studies in the context of WA are limited to the best of our knowledge; the only study in WA established the relationship between cannabis use and opioid misuse in a narrow population and did not assess the relationship between the frequency of cannabis use and opioid-related outcomes.^4,6^ Our study will fill these gaps in the literature through an evaluation of the association between cannabis use and opioid use among adults in WA in 2019 and 2021, allowing us to assess if COVID-19 modified the association and if individuals who use cannabis in varying frequencies for medical use or other use may have higher or lower uptake of opioid. We hypothesized that there would be a higher prevalence of opioid use among participants who reported cannabis use compared to participants who did not report cannabis use and that the association between cannabis and opioid use would be stronger during COVID-19 relative to pre-COVID. Our study will also evaluate the association between the frequency of cannabis use and opioid misuse. We hypothesized that there would be a higher prevalence of opioid misuse among participants who reported a higher frequency of cannabis use compared to participants who reported a lower frequency of cannabis use.

## METHODS

### Study Design and Data Source

We used cross-sectional data from the 2019 and 2021 WA Behavioral Risk Factor Surveillance System (BRFSS) surveys to explore the association between cannabis use and opioid use among adults in WA and the extent to which COVID-19 may have modified this association. BRFSS is an annual telephone survey that collects data on health-related risk behaviors, medical conditions, and individual exposures sampling non-institutionalized adults (18 years of age and older) in the US.^9^ BRFSS uses core questions as well as optional modules and state-added questions that are determined by each state.^9^ In 2019 and 2021, Washington State included state-added questions on cannabis and opioid use.^9^ The University of Washington Institutional Review Board has determined that the use of BRFSS data for research does not involve “human subjects” and therefore requires neither institutional review board review nor an exempt determination.

### Study Population

Our study included respondents to the WA BRFSS who had complete responses to the WA state-added BRFSS questions on cannabis and opioid use. Participants who responded, “Don’t Know,” “Don’t Know/Not Sure,” “Refused,” or had missing responses for questions on cannabis, opioids, or selected confounders were excluded from our analyses.

### Measures

#### Exposure

The primary exposure of interest was current cannabis use and was determined based on the question, “Have you used cannabis in the past 30 days?” Participants who reported “No Use in past 30 Days” were categorized as “no cannabis use.” Participants who reported any use in past 30 days were categorized as “current cannabis use.”

We also considered the frequency of cannabis use, based on the WA state-added BRFSS question, “During the past 30 days, on how many days did you use cannabis or hashish (grass, hash, or pot)?” We used this question to categorize the frequency of cannabis use. Based on previous literature on cannabis use, ^10^ we transformed the numeric responses into four categories: “30” was categorized as “Daily use,” “6-29” was categorized as “Frequent Use,” “1-5” was categorized as “Infrequent use,” and “0” was categorized as “Non-use.”

#### Outcome

The primary outcome of interest was opioid use and was assessed based on the question, “During the past 12 months, did you use prescription pain medication, like Vicodin or OxyContin?” Participants who responded “No” were categorized as “Opioid Non-use.” Participants who responded “Yes” were categorized as “Opioid use,” regardless of frequency or reason for use.

The secondary outcome of interest was opioid misuse and was assessed among those who reported any opioid use in the prior 12 months based on two questions: 1.) “During the past 12 months, did you use a prescription pain medication that was not prescribed specifically to you by a healthcare provider?” and 2.) “During the past 12 months, did you ever use any of the prescription pain medication more often or in higher doses than directed by a healthcare provider?” We created a new dichotomous variable in which participants who responded “Yes’’ to either question were categorized as “Opioid misuse” and participants who responded “No” to both questions were categorized as “Opioid use as prescribed.”

#### Confounders

We selected confounders *a priori* based on prior literature.^4,5,6^ We adjusted for insurance status, sex, age category, race, household income, year of survey, education level, and having at least one comorbidity. Insurance status was based on two variables. In 2019, insurance status was based on a binary variable (Yes/No) that asked the question, “Do you have any kind of health care coverage, including health insurance, prepaid plans such as HMOs, government plans such as Medicare or Indian Health Service?”. In 2021, reported insurance status was based on the question: “What is the current primary source of your health insurance?”. This was calculated into a binary variable (Yes/No) indicating if respondents have health insurance. We combined these into a single variable in which participants who responded “Yes” were categorized as “Insured” and participants who responded “No” were categorized as “Uninsured.” Sex was a binary variable (Male/Female). Age categorized from self-reported age into three groups: “18 to 34,” “35 to 54,” and “55 or older.” Having at least one comorbidity was based on four binary variables (Yes/No) that asked the questions: 1.) “Have you EVER been told by a doctor, nurse, or other health professional that you had any other types of cancer?” 2.) “Have you EVER been told…that you had COPD (Chronic Obstructive Pulmonary Disease), emphysema or chronic bronchitis?” 3.) “Have you EVER been told…that you had angina or coronary heart disease?” 4.) “Have you EVER been told…that you had some form of arthritis, rheumatoid arthritis, gout, lupus, or fibromyalgia?” Individuals who responded “Yes” to any of the four questions were categorized as having “at least one comorbidity.” Individuals who responded “No” to all four questions were categorized as having “no comorbidities.” Race was based on an imputed variable with the categories of “White,” “Black,” “Asian,” “Native American,” “Hispanic,” and “Another Race.” We included those that responded as “Native American” in the category of “Another Race” due to a small sample size (<1%). Household income was based on two variables that asked “What is your annual household income from all sources…?” and then offered a number of categories to choose from. In 2019, categories included “Less than $10,000,” “Less than $15,000,” “Less than $20,000,” “Less than $25,000,” and “Less than $35,000,” “Less than $50,000,” “Less than $100,000,” and “$100,000 or more.” In 2021, all previous responses were included, but the 2019 response “$100,000 or more” was replaced with “Less than $100,000” and included the additional categories of “Less than $150,000,” “Less than $200,000,” and “$200,000 or more.” We combined all of the 2021 categories that had reported incomes over $100,000 to merge these two variables into one new categorical variable with the following categories: “<25,000,” “$25000-$50,000,” “$50,000-$75,000,” “$75,000-100,000,” and “>$100,000.” Education level was determined based on the question, “What is the highest grade or year of school you completed?” Participant responses were assigned to four categories: “Less than High School,” “Graduated High School,” “Technical/AA degree,” and “College Graduate.”

#### Effect Modifiers

We examined whether the survey year (2019/2021) modified the association between cannabis and opioid use. We hypothesized that reported opioid and cannabis uptake may increase following the COVID-19 pandemic due to stressors and healthcare disruptions.

### Statistical Analysis

First, we performed descriptive statistical analyses of our study population. We calculated the weighted percentage for variables of interest to assess the prevalence of covariates among cannabis groups. The BRFSS weighting process is intended to adjust for noncoverage and nonresponse to produce numbers and proportions that equal population estimates, therefore allowing users to generalize from the sample population.

We then conducted a survey-weighted multivariable Poisson regression to generate crude prevalence ratios (PR), adjusted prevalence ratios (aPR), and 95% CIs to estimate the association between cannabis use and any opioid use in WA for the years 2019 and 2021 separately and combined.

Finally, we performed a sensitivity analysis with the Chi-Square test for trend to generate crude prevalence ratios (PR), adjusted prevalence ratios (aPR), and 95% CIs for the association between frequency of cannabis use and opioid misuse in WA.^11,12,13^ All data processing and statistical analyses were performed with R version 4.2.1.^14^

## RESULTS

There were 25,540 respondents to the 2019 (n=12,966) and 2021 (n=12,574) WA BRFSS combined. We excluded 4,615 (18%) participants for missing responses to questions on cannabis use and/or opioid use. Observations were removed when confounding variables had less than 5% unweighted missing data. Our final sample included 20,925 participants to examine the association between cannabis use and opioid use and 2,168 to examine cannabis use and opioid misuse among those with any opioid use (**Figure 3**).

**Figure 3.**
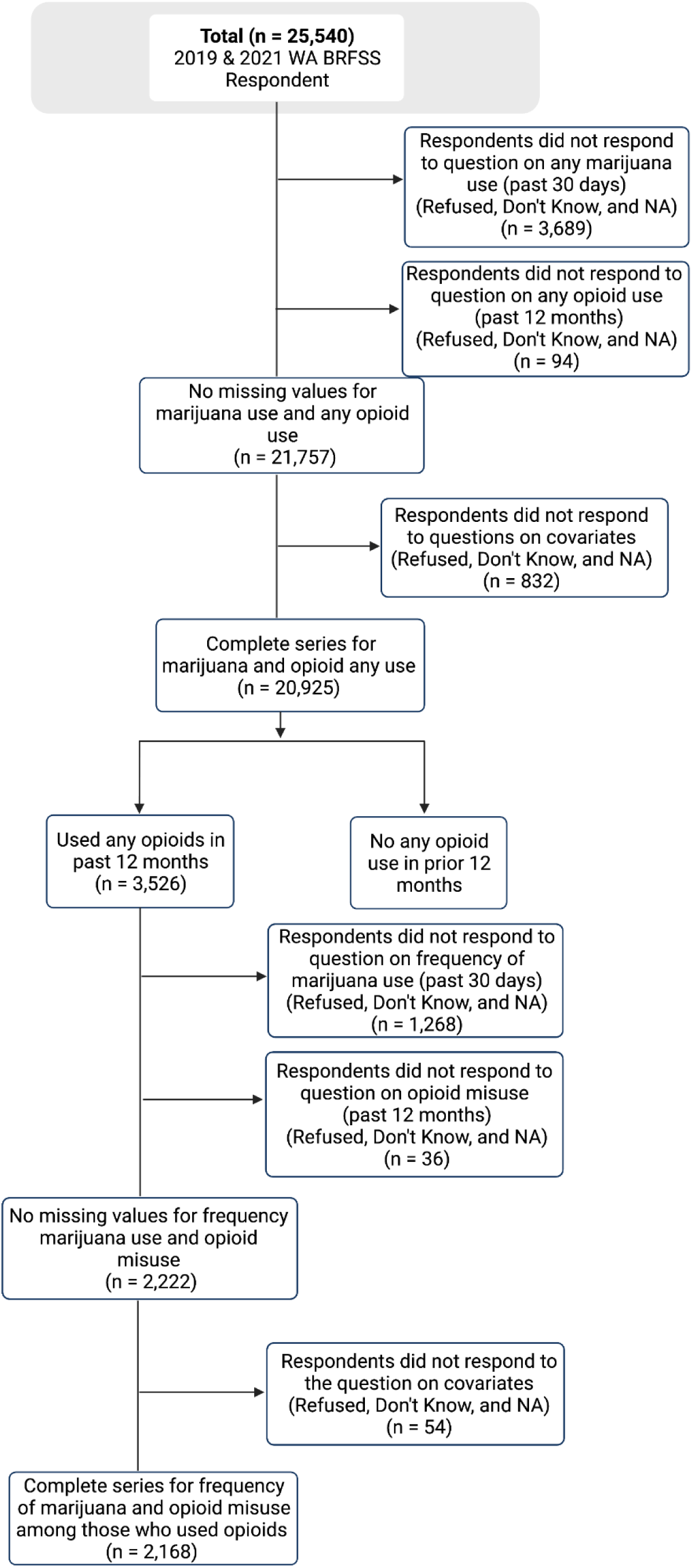
Flowchart illustrating the inclusion and exclusion process of respondents from the 2019 and 2021 Washington Behavioral Risk Factor Surveillance System (WA BRFSS).

The overall prevalence of cannabis use in the past 30 days was 14.4%. A greater proportion of those who used cannabis were younger, white, male, completed some college or a technical degree, insured, and had no comorbidities (**Table 2**).

**Table 1.**
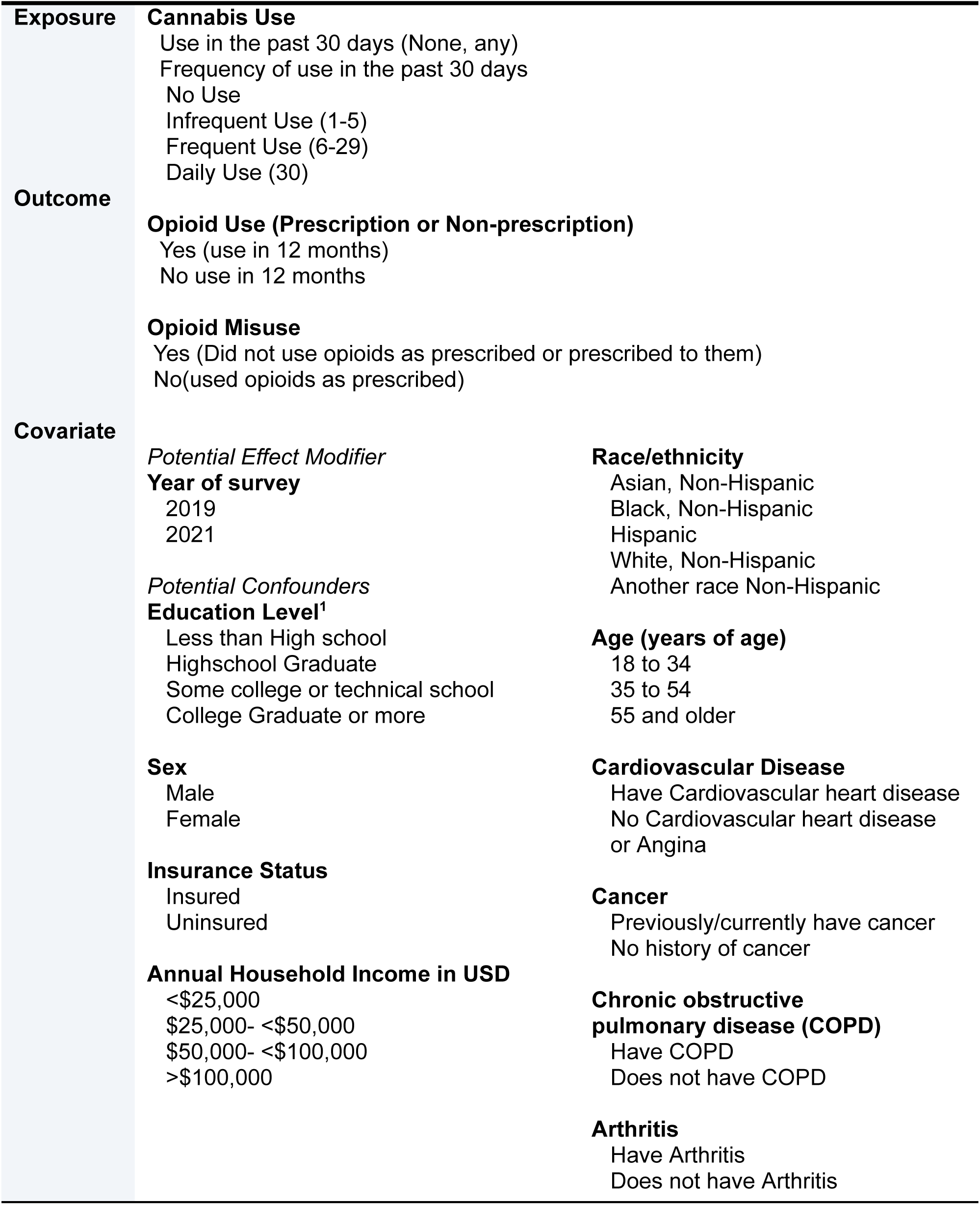
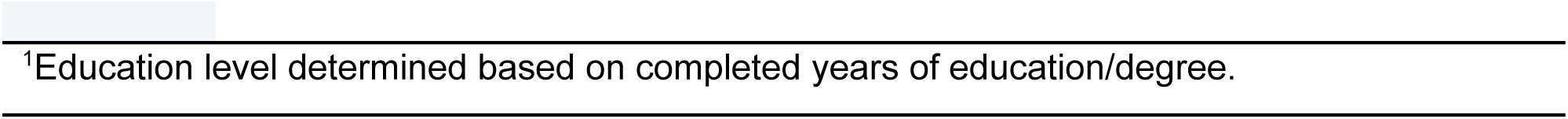
Exposures, outcomes, and potential covariates of interest for the association between cannabis use and opioid use among adults in WA during 2019 and 2021.

**Table 2.**
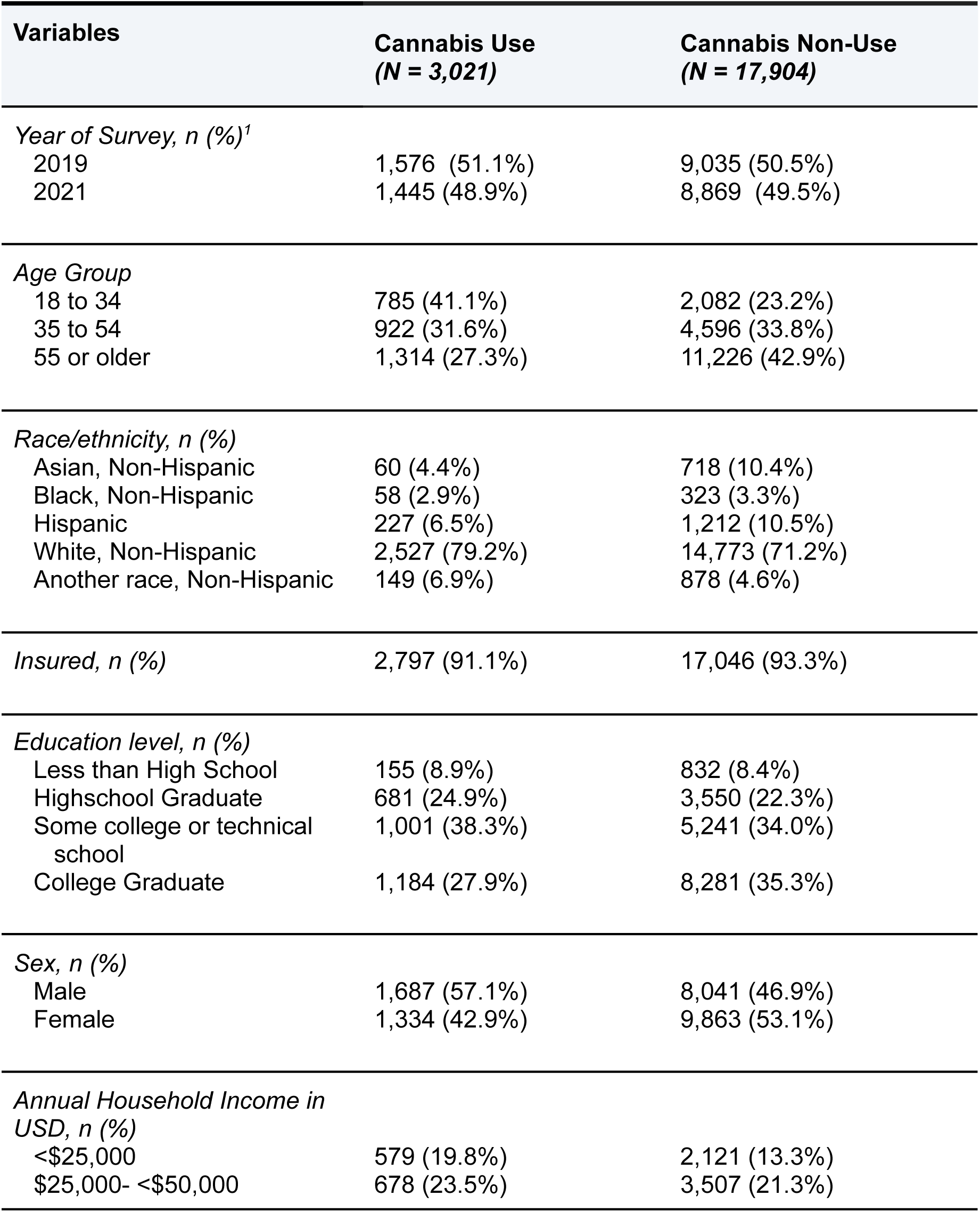
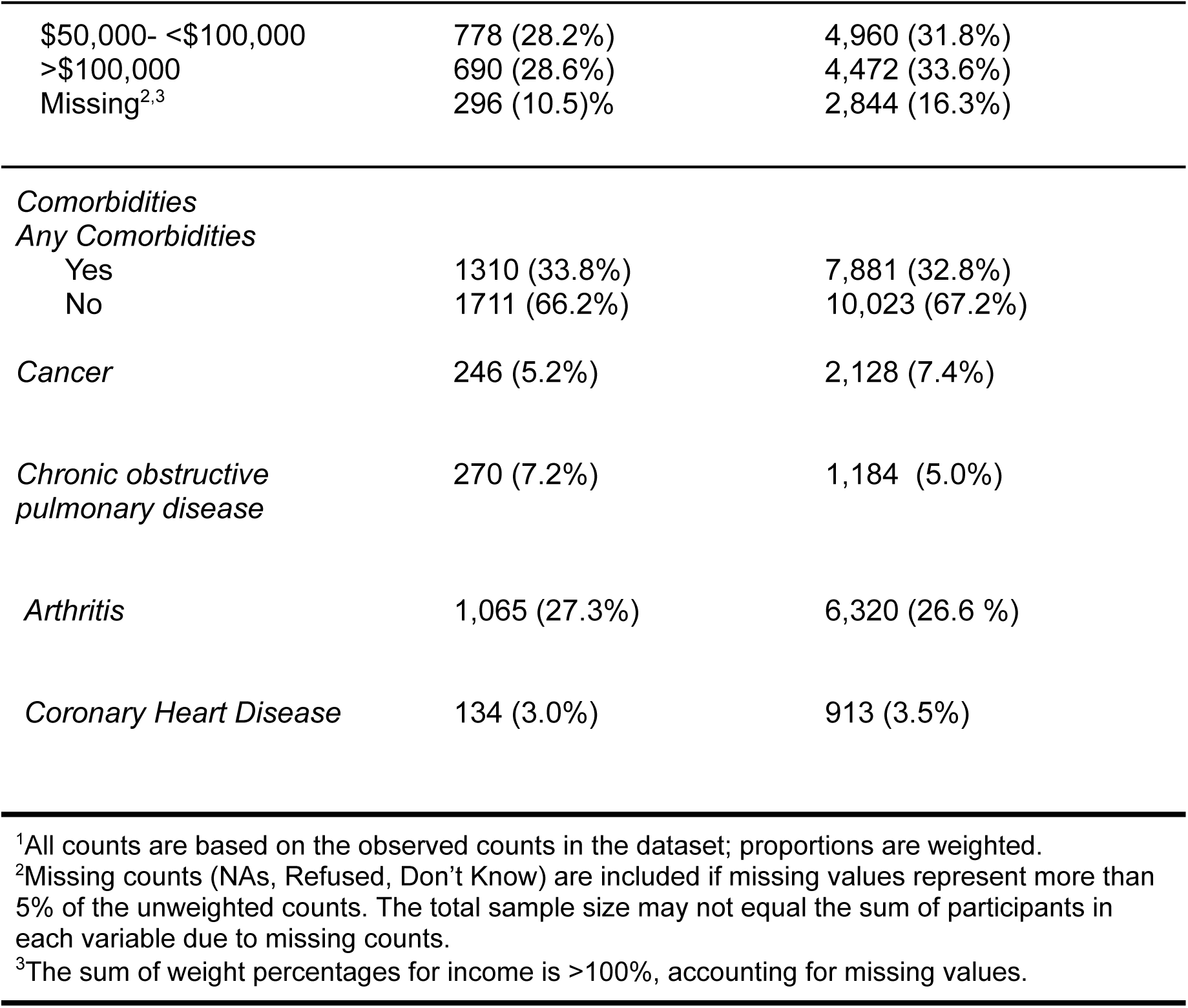
Characteristics of participants by reported cannabis use in the past 30 days in Washington State BRFSS 2019 and 2021 data.

| **Variables** | **Cannabis Use**<br>**(N = 3,021)** | **Cannabis Non-Use**<br>**(N = 17,904)** |
| --- | --- | --- |
| *Year of Survey, n (%)*<sup>1</sup> |  |  |
| 2019 | 1,576 (51.1%) | 9,035 (50.5%) |
| 2021 | 1,445 (48.9%) | 8,869 (49.5%) |
| *Age Group* |  |  |
| 18 to 34 | 785 (41.1%) | 2,082 (23.2%) |
| 35 to 54 | 922 (31.6%) | 4,596 (33.8%) |
| 55 or older | 1,314 (27.3%) | 11,226 (42.9%) |
| *Race/ethnicity, n (%)* |  |  |
| Asian, Non-Hispanic | 60 (4.4%) | 718 (10.4%) |
| Black, Non-Hispanic | 58 (2.9%) | 323 (3.3%) |
| Hispanic | 227 (6.5%) | 1,212 (10.5%) |
| White, Non-Hispanic | 2,527 (79.2%) | 14,773 (71.2%) |
| Another race, Non-Hispanic | 149 (6.9%) | 878 (4.6%) |
| *Insured, n (%)* | 2,797 (91.1%) | 17,046 (93.3%) |
| *Education level, n (%)* |  |  |
| Less than High School | 155 (8.9%) | 832 (8.4%) |
| Highschool Graduate | 681 (24.9%) | 3,550 (22.3%) |
| Some college or technical school | 1,001 (38.3%) | 5,241 (34.0%) |
| College Graduate | 1,184 (27.9%) | 8,281 (35.3%) |
| *Sex, n (%)* |  |  |
| Male | 1,687 (57.1%) | 8,041 (46.9%) |
| Female | 1,334 (42.9%) | 9,863 (53.1%) |
| *Annual Household Income in USD, n (%)* |  |  |
| <\$25,000 | 579 (19.8%) | 2,121 (13.3%) |
| \$25,000- <\$50,000 | 678 (23.5%) | 3,507 (21.3%) |
| \$50,000- <\$100,000 | 778 (28.2%) | 4,960 (31.8%) |
| >\$100,000 | 690 (28.6%) | 4,472 (33.6%) |
| Missing<sup>2,3</sup> | 296 (10.5)% | 2,844 (16.3%) |
| *Comorbidities* |  |  |
| *Any Comorbidities* |  |  |
| Yes | 1310 (33.8%) | 7,881 (32.8%) |
| No | 1711 (66.2%) | 10,023 (67.2%) |
| *Cancer* | 246 (5.2%) | 2,128 (7.4%) |
| *Chronic obstructive pulmonary disease* | 270 (7.2%) | 1,184 (5.0%) |
| *Arthritis* | 1,065 (27.3%) | 6,320 (26.6 %) |
| *Coronary Heart Disease* | 134 (3.0%) | 913 (3.5%) |

<sup>1</sup>All counts are based on the observed counts in the dataset; proportions are weighted.
<sup>2</sup>Missing counts (NAs, Refused, Don't Know) are included if missing values represent more than 5% of the unweighted counts. The total sample size may not equal the sum of participants in each variable due to missing counts.
<sup>3</sup>The sum of weight percentages for income is >100%, accounting for missing values.

The overall weighted prevalence of opioid use in the study population was 13.1%. Among those who used opioids, 20% used cannabis and 80% did not use cannabis. Among participants who reported cannabis use in the past 30 days, the prevalence of opioid use was higher compared to participants who did not report cannabis use in the past 30 days (PR=1.39, 95% CI: 1.25,1.53). This association remained after adjusting for confounders (aPR=1.32, 95% CI: 1.20,1.46) (**Table 3**). The adjusted prevalence ratio between cannabis and opioid use was 1.40 (95% CI: 1.24,1.59) in 2019 and 1.20 (95% CI: 1.01,1.41) in 2021 (**Table 3**).

**Table 3:** Prevalence ratios for the association between cannabis use (past 30 days) and opioid use (past 12 months) overall, and by year (2019 and 2021) in Washington State, Behavioral Risk Factor Surveillance System (n=20,925).

|  | Any Opioid Use in the past 12 months |  |  |  |  |
| --- | --- | --- | --- | --- | --- |
|  | Use<br>N (%)<sup>2</sup> | Non-use<br>N (%)<sup>2</sup> | Total (n) | Crude PR (95%<br>CI) | Adjusted PR<sup>3</sup> (95%<br>CI) |
| **Both Years** |  |  |  |  |  |
| **Cannabis Use in the past 30 days** | 655 (20) | 2,366 (13) | 3,021 | 1.39 (1.25–1.53)* | 1.32 (1.20–1.46)* |
| **Non-use** | 2,692 (80) | 15,212 (87) | 17,904 | 1.0 (Ref) | 1.0 (Ref) |
| **2019\*\*** |  |  |  |  |  |
| **Yes** | 409 (21) | 1,167 (14) | 1,576 | 1.47 (1.30–1.67)* | 1.40 (1.24–1.59)* |
| **No** | 1,569 (79) | 7,466 (86) | 8,995 | 1.0 (Ref) | 1.0 (Ref) |
| **2021\*\*** |  |  |  |  |  |
| **Yes** | 246 (18) | 1,199 (13) | 1,369 | 1.26 (1.06–1.49)* | 1.20 (1.01–1.41)* |
| **No** | 1,123 (82) | 7,746 (87) | 8,869 | 1.0 (Ref) | 1.0 (Ref) |
| <sup>1</sup>Based on any opioid use in the past 12 months<br><sup>2</sup>Unweighted prevalence of cannabis-use participants reporting opioid use<br><sup>3</sup>Estimated using multivariable Poisson regression survey-weighted prevalence ratio (aPR) adjusted for reported insurance status, imputed sex, age category, race, household income, education, year of survey, and having at least one comorbidity<br>*Significant prevalence ratio<br>**Estimated using linear combination, interaction p-value = 0.21 |  |  |  |  |  |

Among those who used opioids, the prevalence of opioid misuse for participants who reported infrequent and frequent use of cannabis was higher compared to participants who reported no cannabis use, but the association was not statistically significant in either crude or adjusted analyses (**Table 4 & Figure 4**). The adjusted prevalence ratios for infrequent use and frequent use were 1.24 (95% CI: 0.68, 2.26) and 1.44 (95% CI: 0.87, 2.37), respectively (**Table 4**). Among participants who reported daily use of cannabis, the prevalence of opioid misuse was higher compared to participants who reported no use (PR=3.00, 95% CI: 1.96, 4.60) (**Table 4**). This finding remained statistically significant after adjusting for confounders (aPR=2.64, 95% CI: 1.71, 4.10) (**Table 4**).

**Table 4:** Prevalence ratios for the association between cannabis use frequency in the past 30 days and opioid misuse in the past 12 months in Washington State, Behavioral Risk Factor Surveillance System (n=2,168).

| Frequency of Cannabis Use (Days) | Opioid Misuse<sup>1,4</sup> (past 12 Months) |  |  |  |
| --- | --- | --- | --- | --- |
|  | Misuse N<sup>2</sup> (%) n=184 | Total | Crude PR (95% CI) | Adjusted PR<sup>3, 5</sup> (95% CI) |
| No Use | 94 (6) | 1515 | 1.0 (Reference) | 1.0 (Reference) |
| Infrequent Use (1-5) | 23 (10) | 223 | 1.28 (0.71-2.34) | 1.24 (0.68–2.26) |
| Frequent Use (6-29) | 27 (12) | 216 | 1.60 (0.98–2.60) | 1.44 (0.87–2.37) |
| Daily Use (30) | 40 (19) | 214 | 3.00 (1.96–4.60)* | 2.64 (1.71–4.10)* |

<sup>1</sup>Based on opioid usage in the past 12 months <sup>2</sup>Unweighted prevalence of individuals reporting opioid misuse for each frequency category <sup>3</sup>Estimated using multivariable Poisson regression survey-weighted prevalence ratio (aPR) adjusted for reported insurance status, imputed sex, age category, race, household income, education, year of survey, and comorbidities <sup>4</sup>Chi-squared Test for trend in proportions statistically significant (p-value < 0.01) <sup>5</sup>Trend test from adjusted trend model (p-value < 0.01) \* p-value < 0.05

## DISCUSSION

In a representative sample of adults in Washington State in 2019 and 2021, the prevalence of opioid use was higher among individuals who used cannabis compared to those who did not use cannabis. This provides support that cannabis use may be a risk factor for prescription opioid use or the reverse.^5,6^ Furthermore, the association between cannabis use and opioid use was similar in 2019 compared to 2021, suggesting that the associations between cannabis and opioid use prior to COVID-19 were similar after.^4,5,6^ We also found that a higher frequency of cannabis use was associated with a higher prevalence of opioid misuse. Daily cannabis use was associated with approximately a three-fold higher prevalence of opioid misuse compared to no cannabis use.

Examining the relationship between cannabis and opioid use in the United States unveils important considerations. Reports from ecological studies indicate a decrease in opioid-related deaths^15^, and the implementation of medical cannabis laws has led to a reduction in opioid prescription patterns.^16^ It is crucial to acknowledge potential misinterpretations when relying solely on ecological studies. Cannabis laws may have increased awareness of opioid misuse leading to a reduction in overdoses rather than misuse as similarly discussed in Olfson et al.^5^ However, this awareness may not relate to an individual-level behavior between cannabis and opioid use. While the majority of adults using cannabis do not subsequently initiate or escalate opioid use, our findings underscore the importance of careful consideration in ongoing policy deliberations. Cannabis regulations promoting the use of cannabis may not be reflected in a reduction in subsequent other drug abuse especially since the individual-level effect of cannabis on opioid outcome is inconclusive.^17^ Furthermore, reports suggest cannabis legalization increases access and thus increases the risk of controlled substance abuse.^18^ Recent modeling studies projected an increase in opioid misuse and overdose in 2025 unless further interventions are implemented.^19^

In the setting of a harm-reduction approach, understanding the relationship between opioid and non-opioid pain management is particularly important. Pre-clinical studies of controlled substances show that chemicals absorbed from heroin and cannabis have similar effects on brain opioid receptors which can become a protective factor or risk factor for opioid use.^20^

This study aimed to contribute to the growing body of evidence regarding the association between cannabis use and opioid misuse. To contextualize our findings, it is important to consider the broader literature on this topic. Several studies, including the 2018 National Epidemiologic Survey on Alcohol and Related Conditions which was a retrospective cohort study of 34,653 adults and the 2014-16 Young Adult Health Survey which involved three cohorts of approximately 2,000 young adults (18-25 years of age), have consistently indicated an increased risk of non-medical opioid use in individuals who use cannabis, both for medical and non-medical purposes with (OR=2.26, 95% CI: 1.86–3.69) and (non-medical OR=4.64, 95% CI: 2.88–7.49; medical OR=1.65, 95% CI: 1.04–2.62) respectively.^5,6^ This suggests that cannabis use may serve as a risk factor for non-medical opioid use. Our study aligns with these findings, as we observed a similar trend in our study of adults in Washington State in 2019 and 2021.^6^ By replicating this association in WA state, our research adds to the evidence that cannabis use may be a risk factor for opioid use in the general population.

In a prospective cohort study with 2,315 participants receiving pharmacological treatment between 2018 and 2020 OUD at treatment clinics in Ontario, Canada, researchers found that cannabis use was not associated with opioid misuse among those who used cannabis in the past 30 days compared to those who did not use cannabis (OR=1.03, 95% CI: 0.87–1.23).^4^ In addition, a higher frequency of cannabis use was found to decrease the odds of opioid use, but this study was limited to individuals with OUD and only included individuals undergoing pharmacological treatment for OUD and therefore is not generalizable to individuals without OUD.^4^ However growing evidence from other population-based studies has reported a positive association between cannabis use and opioid use.^21, 22^

### Limitations

The study has limitations. Due to the cross-sectional design, we cannot establish the direction of the association between cannabis use and opioid use. It is possible that cannabis use makes individuals more susceptible to using opioids, or that individuals with opioid use use cannabis as an alternative or adjunct to opioids.^21,23,24^ The collected data regarding cannabis and opioid consumption relied on self-reported information rather than being validated through urine toxicology, potentially resulting in underestimations. Reported sex is binary (male/female) and does not specify whether it reports sex assigned at birth or current sex. Participants were asked if they used prescription pain medicine, but the question did not specify if it was prescribed to them.

Second, the WA BRFSS survey is conducted using landline telephones or cellphones in Washington State which excludes households without a landline telephone such as unhoused populations that may be affected by this association. We incorporated weights in our study to represent the broader population to the extent that the data would allow.

Third, our data for opioid use and misuse was limited to the years 2019 and 2021. This may underestimate outcomes for those who are exposed to cannabis and/or stressors of COVID-19 which can affect our power to detect significant associations. However, we believe concerns about the different impacts of COVID-19 across individual experiences should be minimal because the WA BRFSS participants were selected randomly, and do not invalidate the significant results of our study.

## Conclusion

This study examines the relationship between cannabis use and opioid use utilizes data after COVID-19 and contributes to the understanding of how the frequency of cannabis use may affect opioid misuse among adults. In addition, our study builds upon the limited research available on the association between cannabis and opioid use, especially in WA. Establishing the temporality of this relationship will be critical to reducing deaths by drug overdose involving opioids, emphasizing the need for careful consideration in policy deliberations regarding cannabis and opioid regulations. Future research efforts should focus on the continued implementation and further development of the WA BRFSS cannabis use and opioids modules and additional questions to further understand behaviors surrounding these two substances.

## Data Availability

All data produced in the present study are available upon reasonable request to the authors and approval to access Washington Department of Health BRFSS database

